# Genome-Wide Polygenic Scores for Common Traits and Psychiatric Disorders Identify Young Children with Risk for Suicides

**DOI:** 10.1101/2020.12.05.20244467

**Authors:** Yoonjung Yoonie Joo, Seo-Yoon Moon, Hee-Hwan Wang, Hyeonjin Kim, Eun-Ji Lee, Seung-Min Jung, Woo-Young Ahn, Incheol Choi, Jae-Won Kim, Jiook Cha

## Abstract

**Background:** Suicide is the leading cause of death in youth worldwide.^1^ Identifying children with high risk for suicide remains challenging.^2^ Here we test the extents to which genome-wide polygenic scores (GPS) for common traits and psychiatric disorders are linked to the risk for suicide in young children.

**Methods:** We constructed GPSs of 24 traits and psychiatric disorders broadly related to suicidality from 8,212 US children with ages of 9 to 10 from the Adolescent Brain Cognitive Development study. We performed multiple logistic regression to test the association between childhood suicidality, defined as suicidal ideation or suicidal attempt, and the GPSs. Machine learning techniques were used to test the predictive utility of the GPSs and other phenotypic outcomes on suicide and suicidal behaviors in the youth.

**Outcomes:** We identified three GPSs significantly associated with childhood suicidality: Attention deficit hyperactivity disorder (ADHD) (P = 2.83×10^−4^; odds ratio (OR) = 1.12, FDR correction), general happiness with belief that own life is meaningful (P = 1.30×10^−3^; OR = 0.89) and autism spectrum disorder (ASD) (P = 1.81×10^−3^; OR = 1.14). Furthermore, the ASD GPS showed significant interaction with ELS such that a greater polygenic score in the presence of a greater ELS has even greater likelihood of suicidality (with active suicidal ideation, P = 1.39×10^−2^, OR = 1.11). In machine learning predictions, the cross validated and optimized model showed an ROC-AUC of 0.72 and accuracy of 0.756 for the hold-out set of overall suicidal ideation prediction, and showed an ROC-AUC of 0.765 and accuracy of 0.750 for the hold-out set of suicidal attempts.

**Interpretation:** Our results show that childhood suicidality is linked to the GPSs for psychiatric disorders, ADHD and ASD, and for a common trait, general happiness, respectively; and that GPSs for ASD and insomnia, respectively, have synergistic effects on suicidality via an interaction with early life stress. By providing the quantitative account of the polygenic and environmental factors of childhood suicidality in a large, representative population, this study shows the potential utility of the GPS in investigation of childhood suicidality for early screening, intervention, and prevention.

## Introduction

In 2020, in every 10 minutes someone in the U.S. died by suicide. In youth, suicide is the leading cause of death worldwide.^1^ Identifying children with high risk for suicide remains challenging with poor predictive validity.^2^ With high estimated heritability of 30% to 55%, suicide is polygenic.^3^ Although recent Genome-Wide Association Studies (GWAS) revealed several genome-wide significant loci,^4,5^ they are not adequtely powered, remain to be replicatedand have yet to offer risk prediction.

Another key outstanding question regarding childhood suicidality is the existence of the gene-by-environment interaction. Early life stress (ELS) or childhood adversity contributes to psychopathology^6^ and suicidality^7^ through the epigenetic mechanism. In fight with childhood suicidality, testing whether ELS and genetic factors together act synergistically on childhood suicidality will provide an insight into the biological pathway, as well as an actionable target for screening and intervention. No literature reports this.

The capability of risk stratification for suicide using genetics and interacting environmental variables would thus help prevent youth suicide. To this end, we test the extents to which genome-wide polygenic scores^8^,9 for common traits and psychiatric disorders, and their interaction with ELS is linked to the risk for suicide in young children.

## Methods

### Study design and participants

We used data from the ABCD (Adolescent Brain and Cognitive Development) study, the nationwide multisite prospective, longitudinal study. The enrolled samples are 11,878 children with ages of 9 to 10 years old in the US across 21 sites recruited between 2015 and 2019. Ethical approval for the study was obtained from Seoul National University IRB. Full details of the study, measures, and samples can be found elsewhere.^10^

### Genotype data

The saliva samples of the participants were collected and genotyped at Rutgers University Cell and DNA Repository using Affymetrix SmokeScreen Array consisting of 733,293 single nucleotide polymorphisms (SNPs). After removing the SNP with genotype call rate <95%, sample call rate <95%, and minor allele frequency (MAF) <1%, raw genotypes were imputed toward 1000G Phase5 reference panel using the Michigan Imputation Server and phased with Eagle v2.4. Additional quality control (QC) process removed SNPs with genotype call rate <95%, Hardy-Weinberg Equilibrium p-value <1E-06, sample missingness >5%, and MAF <1%. We also removed samples with extreme heterozygosity having F coefficient bigger than 3 standard deviation of the population mean. We conducted Principal Component Analysis (PCA) on genetic variants that are pruned for variants in linkage disequilibrium with an r^2^ > 0.25 in a 200kb window. Since ABCD participants reside on a continuum of genetic ancestry as American population (1000Genome super population), rather than distinct population groups, and none of them belongs to either African or East Asian populations, we did not exclude any genetic outliers based on PCA biplot **(Supplementary Figure)**. Proportional identify-by-descent analysis was performed with the same SNP pruning using PLINK^11^ and we removed moderately related individuals (*pihat* > 0.18). After QC, genotype data were available for 8,496 children.

### Construction of Genome-wide Polygenic Scores (GPS)

For GPS generation, we selected 24 target outcomes that are thought to be related to suicidality. These include personality, cognitive, psychological traits, and psychiatric disorders that are known to be broadly related to suicidality: general happiness,^12,13^ insomnia,^14^ depression,^15^ risk behaviors,^16^ risk tolerance, educational attainment,^17,18^ cognitive performance,^16,17^ snoring,^19^ worry,^20^ IQ,^17^ behaviors: cannabis usage,^21,22^ drink per week,^23^ smoker,^24^ Attention deficit hyperactivity disorder (ADHD),^25^ Autism spectrum disorder (ASD),^26^ major depressive disorder (MDD),^27^ schizophrenia,^28^ bipolar disorder,^29^ post-traumatic stress disorder (PTSD).^30^ For general happiness, four kinds of GPS were built and tested for the study: the participants were asked with different mental health questionnaires for measuring the level of subjective well-being, such as (1) two different GWAS of “how happy are you in general”, (2) “how happy are you with your health in general”, and (3) “to what extent do you feel your life to be meaningful”. All the GWAS summary statistics of the aforementioned outcomes are publically available and have been collected for GPS generation.

We performed clumping and pruning of SNPs using Polygenic Risk Score software 2^31^ with a clumping window of 500 kb, clumping r^2^ of 0.2, and no thresholding of p-value significance on the summary statistics since we wanted to fully incorporate the effects of all the SNPs. The polygenic score for each individual was then computed as a sum of their SNPs adjusting for first 10 PCs, with each SNP being weighted by the effect in the discovery samples.^31^

### Outcomes

Suicidal ideation and attempt was derived from the computerized version of Kiddie Schedule for Affective Disorders and Schizophrenia (KSADS).^32,33^ Besides the GPSs and the suicidality outcome variable, the following variables were considered for covariates in logistic regression or the base model for the classification task. For psychopathology, ABCD Parent Child Behavior Checklist; for intelligence, NIH toolbox;^34^ for family environment, Youth Family Environment Scale; and for early life stress, physical and sexual abuse, household challenges such as family substance abuse, family mental illness, family criminals, parental separation or divorce, and violently treated mothers and emotional and physical neglect (a total composite score was used). Demographic variables include sex, age, race/ethnicity, study site, years of parental education, and parental marital status.

### Statistical Analysis

Associations of the GPSs with suicidal variables within the ABCD children were tested using logistic regression with the following covariates: sex, age, site for sample collection, and maternal education level. After QC and GPS construction, the complete dataset of phenotypic outcomes, GPS, and covariate data were available for 8,212 children and used for statistical analysis. To test the extents to which the genome-wide polygenic scores are useful to predict the suicidal risk in children, we trained stacked ensemble models or super learners^35^ based on the several machine learning models, such as lightGBM, xgboost, general linear model (Driverless AI version 1.8, H2O.ai Inc., CA, USA),^36^ and, as a benchmark to the stacked ensemble model, Gradient Boosting Machine (Scikit-learn 0.21; https://scikit-learn.org/). The input variables included the GPSs, sociodemographic, self-reported, and behavioral variables (e.g., psychopathology, cognitive, early life stress). We rationalized that the multitude of the genetic profile would account for the multi-dimensional risk for suicide. For model training and evaluation, we split the data into 80% and 20% in which samples were balanced (bootstrapping without replacement). Within the 80% training data, we conducted five-fold stratified cross validation and optimized hyperparameters in cross validation sets.

## Results

Children with suicidal ideation or attempts showed similar sociodemographic, behavioral, clinical characteristics to those without suicidal ideation or attempts (**Table 1**). Out of the pre-selected 24 GPSs, in suicidal ideation, a greater GPS of ADHD correlated with a greater likelihood (P = 2.83×10^−4^, odds ratio (OR) = 1.12; FDR correction). Likewise, the ADHD GPS significantly correlated with active (P= 9.75×10^−4^, OR=1.14) and passive suicidal ideation (P=6.63×10^−4^, OR=1.13), respectively (**Table 2**). In active suicidal ideation, a greater GPS of ASD correlated with a greater likelihood of suicidal ideation (P= 1.81×10^−3^, OR=1.14). Conversely, in passive suicidal ideation, a less GPS of general happiness correlated with a greater likelihood of suicidal ideation (P = 1.30×10^−3^, OR = 0.89).

**Table 1.** Sociodemographic and clinical outcomes of the participants.

|  | Children with suicidality |  | Control children |  |
| --- | --- | --- | --- | --- |
|  | Count or Mean | Percentage(%) or SD | Count or Mean | Percentage (%) or SD |
| **Female** | 640 | 39.9% | 4822 | 48.8% |
| **Age (in months)** | 118.8 | 7.54 | 119.0 | 7.45 |
| **Income (bracket in 1~10 scales)** | 7.07 | 2.38 | 7.26 | 2.41 |
| **Marital status of the family (Currently married)** | 998 | 62.3% | 6476 | 65.5% |
| **Mother's EA** | 16.62 | 2.63 | 16.61 | 2.78 |

**Table 2.** Significant association of GPS with Childhood Suicidality the ABCD children (n=8,212) having FDR significance < 0.05.

| Outcome | GPS predictor | OR | 95% OR | P-value | N of cases |
| --- | --- | --- | --- | --- | --- |
|  |  |  |  | (FDR corrected) |  |
| Suicidal ideation | ADHD | 1.123 | (1.06 - 1.20) | 2.83E-04 (2.27E-02) | 1155 |
| Passive suicidal ideation | ADHD | 1.126 | (1.05 - 1.20) | 6.63E-04 (3.11E-02) | 946 |
| Active suicidal ideation | ADHD | 1.15 | (1.06 - 1.25) | 9.75E-04 (3.11E-02) | 611 |
| Passive suicidal ideation | GENERAL HAPPINESS MEANINGFUL | 0.895 | (0.84 - 0.96) | 1.30E-03 (3.11E-02) | 946 |
| Active suicidal ideation | ASD | 1.141 | (1.05 - 1.24) | 1.81E-03 (3.48E-02) | 611 |

Of those three GPSs showing significant correlations with suicidality, we further tested an interaction with ELS. We found a significant ASD-GPS-by-ELS interaction on active suicidal ideation: a greater ASD GPS in the presence of ELS correlated with a synergistic increase in these likelihood of suicidality (P=1.39×10^−2^, OR=1.11; FDR correction for the three analyses). Effects were adjusted for the covariates. No significant associations were found between ELS and suicidality (Ps > 0.24). When stratified by sex, no significant associations were found.

We tested whether the multiple GPSs contributed to prediction of childhood suicidality using machine learning. The following input features were used: the GPSs, socio-demographic (sex, age, marital status, parental education, ethnicity, study site), psychological (Child behavior checklist), cognitive phenotypic (NIH toolbox), family environment (Youth Family Environment Scale), and ELS. In predicting suicidal ideation, the cross-validated and optimized stacked ensemble model showed an ROC-AUC of 0.720, accuracy of 0.756, sensitivity of 0.837, specificity of 0.058, positive predictive value of 0.538, negative predictive value of 0.214 in the balanced held-out test set (N= 476; bootstrap samples) (**Figure 2**). Adding the GPS to the phenotype model resulted in a 2.4% boost in ROC-AUC. In predicting suicidal attempts, the cross validated and optimized stacked ensemble model showed an ROC-AUC of 0.765, accuracy of 0.750, sensitivity of 0.583, specificity of 0.25, positive predictive value of 0.70, negative predictive value of 0.167 in the balanced held-out test set (N= 32; bootstrap samples) (**Figure 2**). Adding the GPS to the phenotype model resulted in a 3.2% boost in ROC-AUC. The stacked ensemble models always outperformed the benchmark Gradient Boosting Machine model.

**Figure 1.**
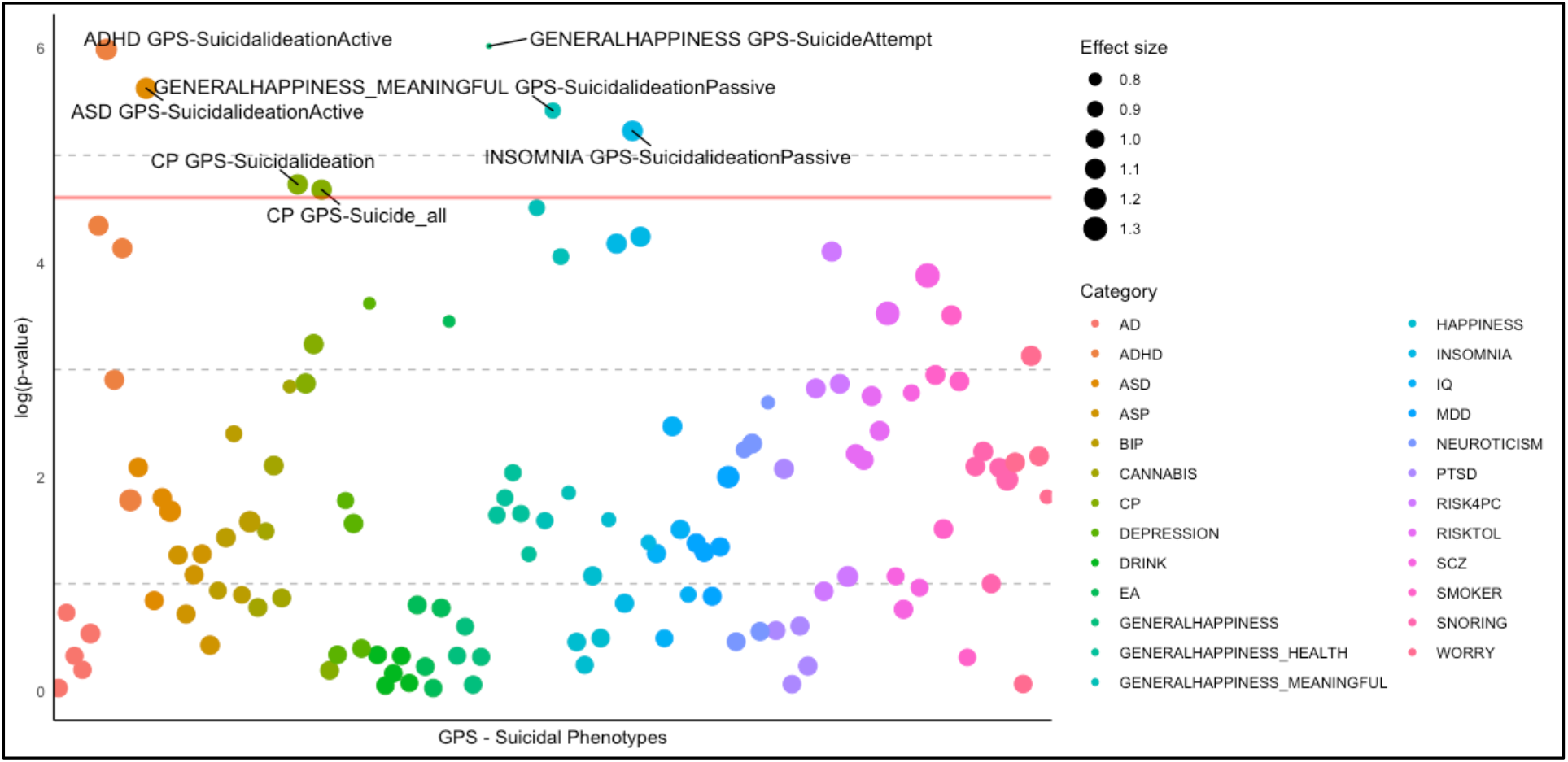
Manhattan plot showing associations of multiple genome-wide polygenic scores of cognitive, psychological, personality traits, and psychiatric disorders with childhood suicidality. Red line indicates FDR corrected P of 0.05. Target outcomes of the GPSs are color-coded. An effect size of each GPS is denoted with the circle size.

**Figure 2.**
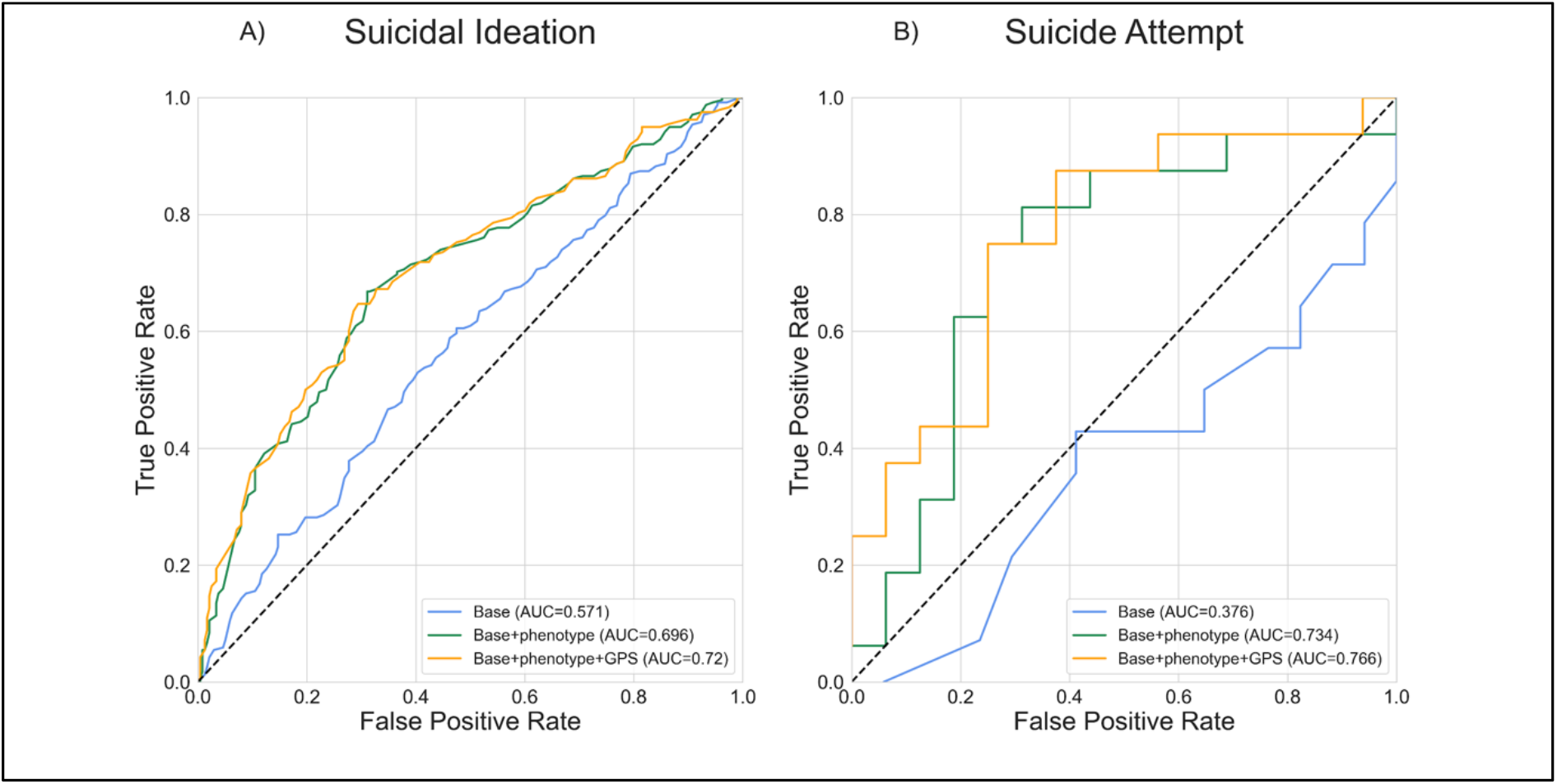
Prediction performance of the machine learning models based on GPS and cognitive, psychological, behavioral, environmental, familial variables. Receiver-Operator Characteristics curves of the models predicting suicidal ideation (A) and suicide attempt (B).

## Discussion

We tested the utility of the GPS in identifying children with risk for suicide. Using large-scale, nation-wide, representative children samples, we found novel associations between childhood suicidality and the GPS of ADHD, ASD -- positive associations -- and general happiness -- a negative association. We next found significant gene-by-environment interactions between the GPS of ASD and ELS,^6^ a known risk factor for childhood suicidality, together acting synergistically on childhood suicidality. In our data-driven predictive modeling, together with the self-reported questionnaires for psychopathology (e.g., CBCL), intelligence, family environment, and socio-demographic variables, inclusion of the multiple GPSs permitted to classify children with suicidality with moderate accuracy. To sum, this study sheds light on the genetic approach to childhood suicidality for better understanding of the etiologic pathway and for prevention and intervention.

Using the multi-GPS method we discovered the association of childhood suicidality with the genetic profiles of psychiatric disorders, i.e., ADHD and ASD, particularly relevant to childhood psychopathology, and of a common trait, i.e., general happiness. For ADHD and ASD, a greater polygenic score correlates with a greater likelihood of suicide, whereas for general happniness, a smaller polygenic score correlates with a greater likelihood of suicide. This is in line with the literature of youth suicide. ADHD in adults has been implicated in completed suicide,^37^ and in youth in suicidal attempts.^38^ Our GPS results may provide the genomic evidence of the link between ADHD and childhood suicidality. The explained variance (McFadden’s pseudo-R^2^)^39^ of suicidal phenotypes by ADHD GPS was approximately 1.5% in children. This estimation is higher than the results from past PRS studies of suicidality, which showed maximum variance explained by 0.13-0.20% with self-harm behaviors^40^ or up to 0.30-0.70% of the phenotypic variance for suicide attempt explained by depression-based GPS.^41,42^

The GPS of ASD not only correlated with suicidal ideation, but also showed a significant interaction with ELS. These results corroborate previous findings of children with ASD being at an increased risk for suicidality.^43,44^ One study shows that children with ASD are 28 times more likely to exhibit suicidal thoughts or behaviors than that of typical counterparts.^45^ This strong link between risk of suicidality and elevated autistic traits could be explained by specific behavioral attributes of both phenotypes, such as poor socialization and problem-solving skill, or increased levels of anxiety.^44^ Although literature has reported several social risk factors underlying the ASD-suicidality link,^45,46^ the genetic and environmental contribution to the overlap of ASD and suicidal behaviors remained unknown.^47^ Our results fill this gap by presenting the shared genetic factors for ASD and suicidality, also in environmental (childhood experiences) dimensions. The results thus further suggest that protective, as opposed to adverse, childhood experiences are particularly important for those with genetic risk of ASD. This might be a potential actionable target for intervention in that population.

In line with our findings, previous literature has reported high suicidal risk in individuals who in addition to ASD also have ADHD^47^. As 40-50% of individuals with ASD are known to have comorbid ADHD,^48^ our significant results of ASD and ADHD GPSs associated with suicidality suggest the genetic liability of psychiatric comorbidity for high suicidal risk in children.

The association of a less GPS of general happiness (believing that her or his life is meaningful) and a greater likelihood of childhood suicidal ideation is novel. Of several measures of subjective well-being, only the belief of own meaningful life is significantly linked to childhood suicidality. Prior studies report a negative correlation between subjective happiness and suicide.^12,13^. Our results corroborate the link by showing genetic underpinning of the relationship.

Our study contributes in several ways to our genomic understanding of suicidal phenotypes and provides a basis for utilizing multiple GPSs for suicidal prediction. The combination of the non-suicidal GPSs, behavioral, psychological scales, and ELS allowed accurate prediction of suicidality with a ROC-AUC up to 0.720 for suicidal ideation and 0.765 for suicidal attempts on the balanced test set. Several previous studies have tested different methods for suicidal risk prediction and showed substantial classification ability having AUC ranging from 0.74 to 0.88, using known sociodemographic and psychiatric risk factors.^49–52^ Although quite successful, some of the models included long-term measurement of risk indices such as 12-month or lifelong risk factors, which are not readily measured in naturalistic clinical practices. Our models showed relatively lower performances than previous studies, but our methodology is promising in that they incorporated short-term or cross-sectional factors that could be accessible and readily measured in clinical settings, on top of the genetic factors. Despite the acceptable accuracy and sensitivity, however, there is much room for improvement regarding specificity and negative predictive value of the model.

Despite the potential utility of the GPS in childhood suicide research discovered here, towards the practical utility in early screening further methodological improvement is needed. Firstly in generating the GPS, searching for the optimal hyper-parameters (e.g.,p-value cutoff or linkage disequilibrium criteria) may lead to less training errors and an improved predictive power. Secondly, future research should find an optimal selection of genetic and environmental risk factors to accurately predict childhood suicidality to prevent suicidal behaviors in youth from the clinical context.

By providing the first quantitative account of the polygenic and environmental factors of childhood suicidality in a large, representative population, this study shows the importance and potential utility of the GPS approach in childhood suicidality in early screening. These findings motivate the future investigation of more effective screening and intervention strategies for children suicidality.

## Data Availability

Codes and data is freely available for reproducibility.

https://github.com/Transconnectome/ConnectomeLab/blob/master/suicide

## Author contributions

Study concept and design: Y.J., J.C.

Acquisition, analysis, or interpretation of data: Y.J., H.K., S.M., H.W.

Drafting of the manuscript: Y.J., S.M., H.W., J.C.

Critical revision of the manuscript for important intellectual content: Y.J., J.K., I.C., J.C., W.-Y.A.

Statistical analysis: Y.J., S.M., H.W.

Obtained funding: I.C., J.C.

Study supervision: J.C.

## Data and Code Availability

Codes and data is freely available for reproducibility (https://github.com/Transconnectome/ConnectomeLab/blob/master/suicide).

## Acknowledgement

This work was supported by the New Faculty Startup Fund from Seoul National University, Research grant from the Center for Happiness via the Center for Social Sciences at Seoul National University at Seoul National University, and the H2O.ai Academic Program at H2O inc., CA, USA.

## Supplementary Materials

**Supplementary Figure 1.**
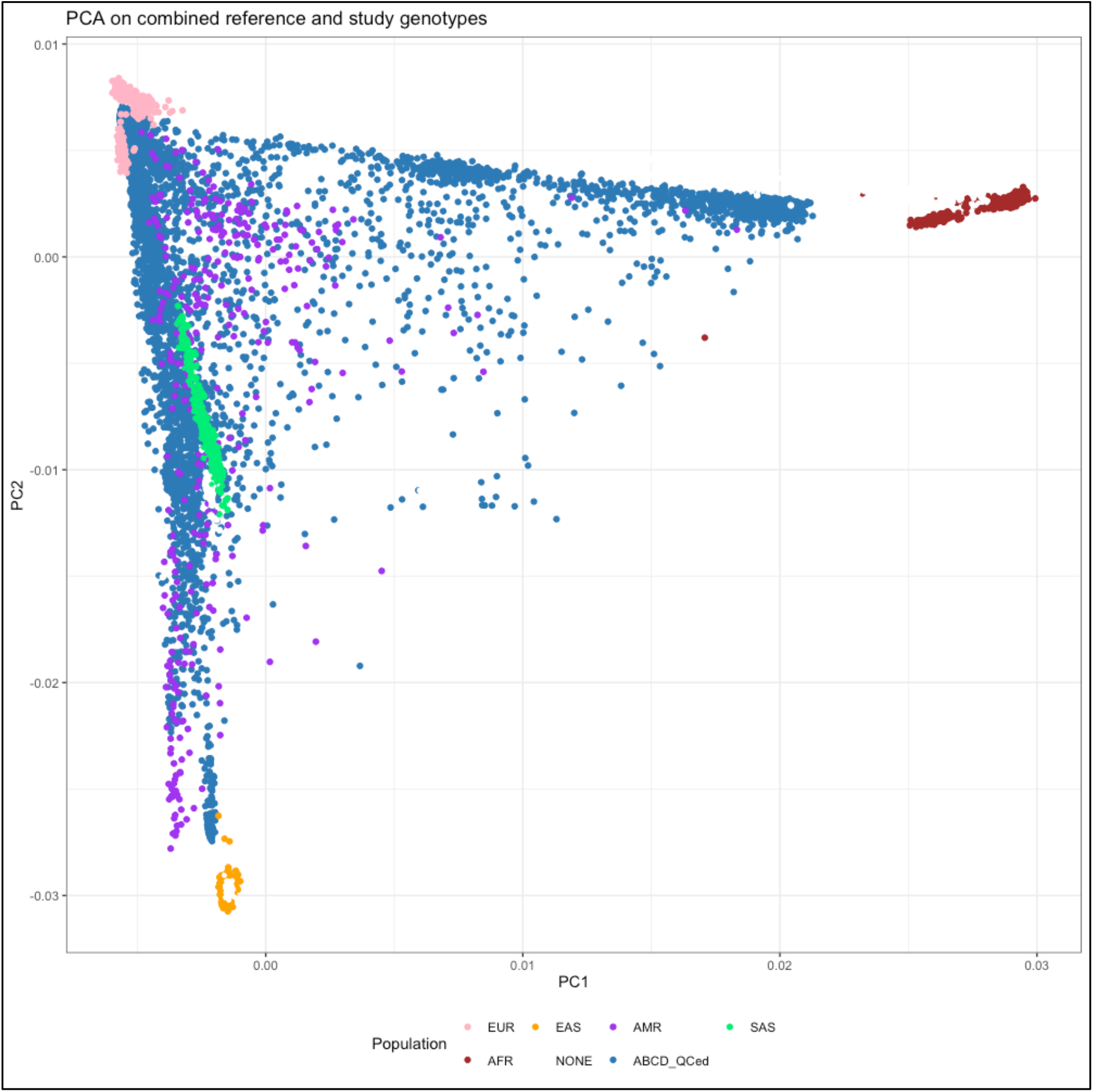
Biplot from Principal Component Analysis on the combined genotype data of 1000Genome reference panel^1^ and the ABCD study participants. (colored blue). Super-populations are defined broadly as: AFR African, AMR admixed American, EAS East Asian, EUR European, SAS South Asian. To facilitate the understanding of genetic ancestry of the study participants, PCA was performed with 1000Genome phase3 reference panel and plotted with the first two principal components. Since ABCD participants reside on a continuum of genetic ancestry as American population (AMR), rather than distinct population groups, and none of them belongs to either African (AFR) or East Asian (EAS) populations, we did not exclude any genetic outliers based on the plot.

**Supplementary table 1.**
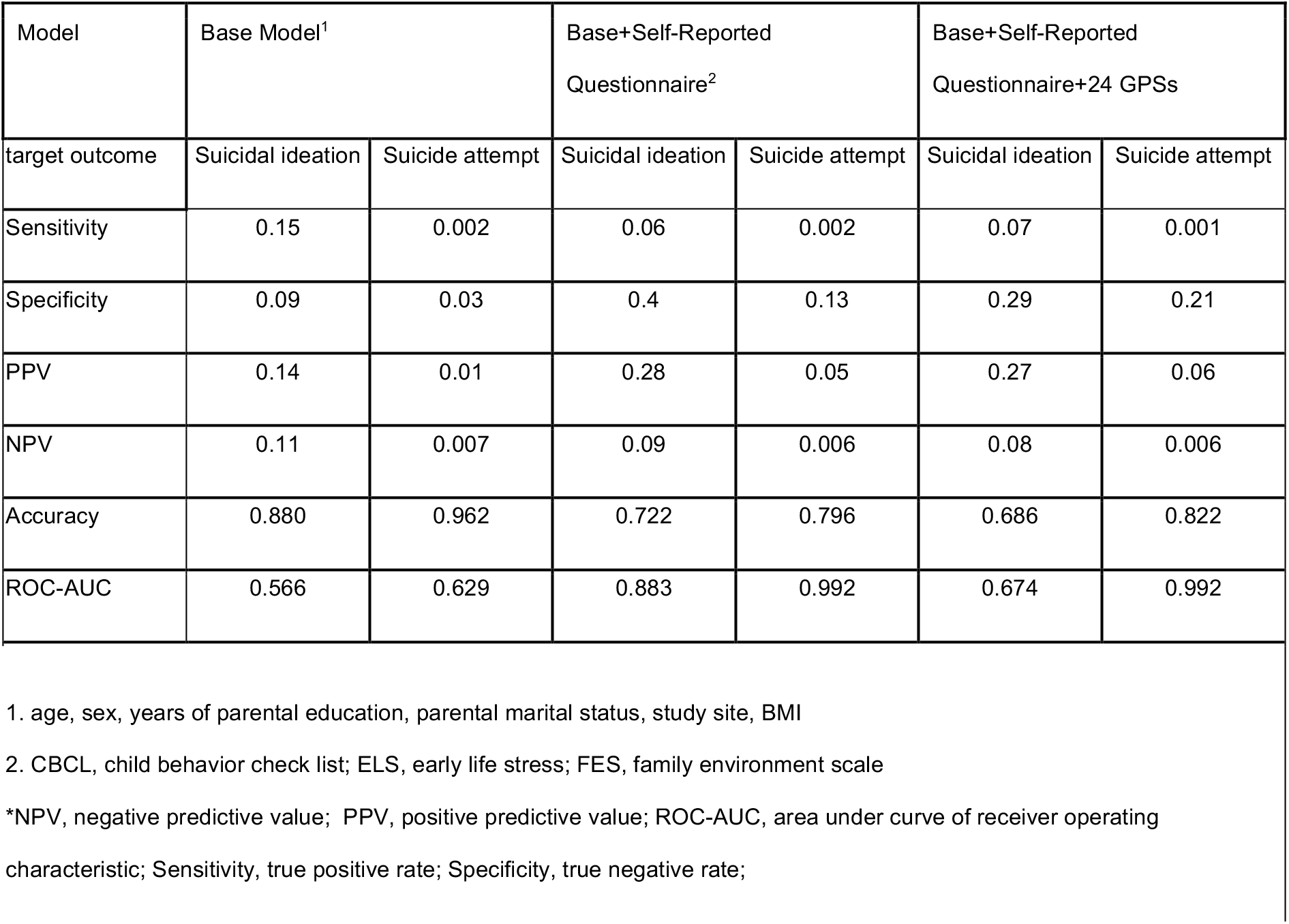
Performance of suicidality classification models in the imbalanced validation set (in suicidal ideation, 956 cases and 7,030 controls; in suicide attempt, 64 cases and 8,366 controls)

| Model | Base Model<sup>1</sup> |  | Base+Self-Reported Questionnaire<sup>2</sup> |  | Base+Self-Reported Questionnaire+24 GPSs |  |
| --- | --- | --- | --- | --- | --- | --- |
|  | Suicidal ideation | Suicide attempt | Suicidal ideation | Suicide attempt | Suicidal ideation | Suicide attempt |
| Sensitivity | 0.15 | 0.002 | 0.06 | 0.002 | 0.07 | 0.001 |
| Specificity | 0.09 | 0.03 | 0.4 | 0.13 | 0.29 | 0.21 |
| PPV | 0.14 | 0.01 | 0.28 | 0.05 | 0.27 | 0.06 |
| NPV | 0.11 | 0.007 | 0.09 | 0.006 | 0.08 | 0.006 |
| Accuracy | 0.880 | 0.962 | 0.722 | 0.796 | 0.686 | 0.822 |
| ROC-AUC | 0.566 | 0.629 | 0.883 | 0.992 | 0.674 | 0.992 |

1. age, sex, years of parental education, parental marital status, study site, BMI
2. CBCL, child behavior check list; ELS, early life stress; FES, family environment scale
\*NPV, negative predictive value; PPV, positive predictive value; ROC-AUC, area under curve of receiver operating characteristic; Sensitivity, true positive rate; Specificity, true negative rate;

**Supplementary table 2.** Performance of suicidality classification models in the balanced test set (in suicidal ideation, 238 cases and 238 controls; in suicide attempt, 16 cases and 16 controls)

| Model | Base Model<sup>1</sup> |  | Base+Self-Reported Questionnaire<sup>2</sup> |  | Base+Self-Reported Questionnaire+24 GPSs |  |
| --- | --- | --- | --- | --- | --- | --- |
| target outcome | Suicidal ideation | Suicide attempt | Suicidal ideation | Suicide attempt | Suicidal ideation | Suicide attempt |
| Sensitivity | 0.96 | 1 | 0.53 | 0.54 | 0.837 | 0.583 |
| Specificity | 0 | 0 | 0.45 | 0.38 | 0.058 | 0.25 |
| PPV | 0.5 | 0.45 | 0.66 | 0.72 | 0.538 | 0.7 |
| NPV | 0.15 | 0 | 0.32 | 0.21 | 0.214 | 0.167 |
| Accuracy | 0.565 | 0.516 | 0.679 | 0.75 | 0.756 | 0.75 |
| AUC | 0.571 | 0.376 | 0.695 | 0.734 | 0.72 | 0.766 |

1. age, sex, years of parental education, parental marital status, study site, BMI
2. CBCL, child behavior check list; ELS, early life stress; FES, family environment scale
\*NPV, negative predictive value; PPV, positive predictive value; ROC-AUC, area under curve of receiver operating characteristic; Sensitivity, true positive rate; Specificity, true negative rate;

## References

1. Cunningham RM, Walton MA, Carter PM. The Major Causes of Death in Children and Adolescents in the United States. New England Journal of Medicine. 2018;379(25):2468–2475. doi:10.1056/nejmsr1804754

2. Belsher BE, Smolenski DJ, Pruitt LD, et al. Prediction Models for Suicide Attempts and Deaths: A Systematic Review and Simulation. JAMA Psychiatry. 2019;76(6):642–651.

3. Sokolowski M, Wasserman J, Wasserman D. Polygenic associations of neurodevelopmental genes in suicide attempt. Mol Psychiatry. 2016;21(10):1381–1390.

4. Mullins N, Bigdeli TB, Børglum AD, et al. GWAS of Suicide Attempt in Psychiatric Disorders and Association With Major Depression Polygenic Risk Scores. Am J Psychiatry. 2019;176(8):651–660.

5. Otsuka I, Akiyama M, Shirakawa O, et al. Genome-wide association studies identify polygenic effects for completed suicide in the Japanese population. Neuropsychopharmacology. 2019;44(12):2119–2124.

6. Turecki G, Ota VK, Belangero SI, Jackowski A, Kaufman J. Early life adversity, genomic plasticity, and psychopathology. Lancet Psychiatry. 2014;1(6):461–466.

7. Turecki G, Brent DA. Suicide and suicidal behaviour. The Lancet. 2016;387(10024):1227–1239. doi:10.1016/s0140-6736(15)00234-2

8. Hunter DJ, Longo DL. The Precision of Evidence Needed to Practice “Precision Medicine.” New England Journal of Medicine. 2019;380(25):2472–2474. doi:10.1056/nejme1906088

9. Sugrue LP, Desikan RS. What Are Polygenic Scores and Why Are They Important? JAMA. 2019;321(18):1820–1821.

10. Karcher NR, Barch DM. The ABCD study: understanding the development of risk for mental and physical health outcomes. Neuropsychopharmacology. Published online June 15, 2020. doi:10.1038/s41386-020-0736-6

11. Chang CC, Chow CC, Tellier LC, Vattikuti S, Purcell SM, Lee JJ. Second-generation PLINK: rising to the challenge of larger and richer datasets. Gigascience. 2015;4:7.

12. Pompili M, Innamorati M, Lamis DA, et al. The Interplay Between Suicide Risk, Cognitive Vulnerability, Subjective Happiness and Depression Among Students. Current Psychology. 2016;35(3):450–458. doi:10.1007/s12144-015-9313-2

13. Hsu C-Y, Chang S-S, Yip PSF. Subjective wellbeing, suicide and socioeconomic factors: an ecological analysis in Hong Kong. Epidemiology and Psychiatric Sciences. 2019;28(1):112–130. doi:10.1017/s2045796018000124

14. Hamilton JL, Buysse DJ. Reducing Suicidality Through Insomnia Treatment: Critical Next Steps in Suicide Prevention. American Journal of Psychiatry. 2019;176(11):897–899. doi:10.1176/appi.ajp.2019.19080888

15. Gabrielle A. Carlson DPC. Suicidal Behavior and Depression in Children and Adolescents. J Am Acad Child Psychiatry. 1982;21(4):361–368.

16. King RA, Schwab-Stone M, Flisher AJ, et al. Psychosocial and risk behavior correlates of youth suicide attempts and suicidal ideation. J Am Acad Child Adolesc Psychiatry. 2001;40(7):837–846.

17. Keilp JG, Sackeim HA, Brodsky BS, Oquendo MA, Malone KM, Mann JJ. Neuropsychological dysfunction in depressed suicide attempters. Am J Psychiatry. 2001;158(5):735–741.

18. Rosoff DB, Kaminsky ZA, McIntosh AM, Davey Smith G, Lohoff FW. Educational attainment reduces the risk of suicide attempt among individuals with and without psychiatric disorders independent of cognition: a bidirectional and multivariable Mendelian randomization study with more than 815,000 participants. Transl Psychiatry. 2020;10(1):388.

19. Becker SP, Dvorsky MR, Holdaway AS, Luebbe AM. Sleep problems and suicidal behaviors in college students. J Psychiatr Res. 2018;99:122–128.

20. Gorday JY, Rogers ML, Joiner TE. Examining characteristics of worry in relation to depression, anxiety, and suicidal ideation and attempts. J Psychiatr Res. 2018;107:97–103.

21. Price C, Hemmingsson T, Lewis G, Zammit S, Allebeck P. Cannabis and suicide: longitudinal study. Br J Psychiatry. 2009;195(6):492–497.

22. Orri M, Séguin JR, Castellanos-Ryan N, et al. Agenetically informed study on the association of cannabis, alcohol, and tobacco smoking with suicide attempt. Molecular Psychiatry. Published online 2020. doi:10.1038/s41380-020-0785-6

23. Borges G, Bagge CL, Cherpitel CJ, Conner KR, Orozco R, Rossow I. A meta-analysis of acute use of alcohol and the risk of suicide attempt. Psychol Med. 2017;47(5):949–957.

24. Green M, Turner S, Sareen J. Smoking and suicide: biological and social evidence and causal mechanisms. J Epidemiol Community Health. 2017;71(9):839–840.

25. Beauchaine TP, Ben-David I, Bos M. ADHD, financial distress, and suicide in adulthood: A population study. Sci Adv. 2020;6(40). doi:10.1126/sciadv.aba1551

26. Cassidy S, Rodgers J. Understanding and prevention of suicide in autism. Lancet Psychiatry. 2017;4(6):e11.

27. Angst J, Angst F, Stassen HH. Suicide risk in patients with major depressive disorder. J Clin Psychiatry. 1999;60 Suppl 2:57-62; discussion 75-76, 113-116.

28. Palmer BA, Shane Pankratz V, Bostwick JM. The Lifetime Risk of Suicide in Schizophrenia. Archives of General Psychiatry. 2005;62(3):247. doi:10.1001/archpsyc.62.3.247

29. Hawton K, Sutton L, Haw C, Sinclair J, Harriss L. Suicide and attempted suicide in bipolar disorder: a systematic review of risk factors. J Clin Psychiatry. 2005;66(6):693–704.

30. Gradus JL, Qin P, Lincoln AK, et al. Posttraumatic Stress Disorder and Completed Suicide. American Journal of Epidemiology. 2010;171(6):721–727. doi:10.1093/aje/kwp456

31. Choi SW, Mak TS-H, O’Reilly PF. Tutorial: a guide to performing polygenic risk score analyses. Nat Protoc. 2020;15(9):2759–2772.

32. Kaufman J, Birmaher B, Brent D, et al. Schedule for Affective Disorders and Schizophrenia for School-Age Children-Present and Lifetime Version (K-SADS-PL): initial reliability and validity data. J Am Acad Child Adolesc Psychiatry. 1997;36(7):980–988.

33. Kaufman J, Townsend LD, Kobak K. The Computerized Kiddie Schedule for Affective Disorders and Schizophrenia (KSADS): Development and Administration Guidelines. Journal of the American Academy of Child & Adolescent Psychiatry. 2017;56(10):S357. doi:10.1016/j.jaac.2017.07.770

34. Luciana M, Bjork JM, Nagel BJ, et al. Adolescent neurocognitive development and impacts of substance use: Overview of the adolescent brain cognitive development (ABCD) baseline neurocognition battery. Dev Cogn Neurosci. 2018;32:67–79.

35. Laan MJ van der, van der Laan MJ, Polley EC, Hubbard AE. Super Learner. Statistical Applications in Genetics and Molecular Biology. 2007;6(1). doi:10.2202/1544-6115.1309

36. H2O Driverless AI - Open Source Leader in AI and ML. Accessed December 1, 2020. https://www.h2o.ai/products/h2o-driverless-ai/

37. James A, Lai FH, Dahl C. Attention deficit hyperactivity disorder and suicide: a review of possible associations. Acta Psychiatr Scand. 2004;110(6):408–415.

38. Chronis-Tuscano A, Molina BSG, Pelham WE, et al. Very early predictors of adolescent depression and suicide attempts in children with attention-deficit/hyperactivity disorder. Arch Gen Psychiatry. 2010;67(10):1044–1051.

39. McFadden D. Conditional Logit Analysis of Qualitative Choice Behavior., 1973.

40. Campos AI, Verweij KJH, Statham DJ, et al. Genetic aetiology of self-harm ideation and behaviour. Sci Rep. 2020;10(1):9713.

41. Levey DF, Polimanti R, Cheng Z, et al. Genetic associations with suicide attempt severity and genetic overlap with major depression. Transl Psychiatry. 2019;9(1):22.

42. Shen H, Gelaye B, Huang H, Rondon MB, Sanchez S, Duncan LE. Polygenic prediction and GWAS of depression, PTSD, and suicidal ideation/self-harm in a Peruvian cohort. Neuropsychopharmacology. 2020;45(10):1595–1602. doi:10.1038/s41386-020-0603-5

43. Howe SJ, Hewitt K, Baraskewich J, Cassidy S, McMorris CA. Suicidality Among Children and Youth With and Without Autism Spectrum Disorder: A Systematic Review of Existing Risk Assessment Tools. J Autism Dev Disord. 2020;50(10):3462–3476.

44. Chen Y-Y, Chen Y-L, Gau SS-F. Suicidality in Children with Elevated Autistic Traits. Autism Res. 2020;13(10):1811–1821.

45. Mayes SD, Gorman AA, Hillwig-Garcia J, Syed E. Suicide ideation and attempts in children with autism. Research in Autism Spectrum Disorders. 2013;7(1):109–119. doi:10.1016/j.rasd.2012.07.009

46. McDonnell CG, DeLucia EA, Hayden EP, et al. An Exploratory Analysis of Predictors of Youth Suicide-Related Behaviors in Autism Spectrum Disorder: Implications for Prevention Science. J Autism Dev Disord. 2020;50(10):3531–3544.

47. Hirvikoski T, Boman M, Chen Q, et al. Individual risk and familial liability for suicide attempt and suicide in autism: a population-based study. Psychol Med. 2020;50(9):1463–1474.

48. Ljung T, Chen Q, Lichtenstein P, Larsson H. Common etiological factors of attention-deficit/hyperactivity disorder and suicidal behavior: a population-based study in Sweden. JAMA Psychiatry. 2014;71(8):958–964.

49. Borges G, Nock MK, Haro Abad JM, et al. Twelve-month prevalence of and risk factors for suicide attempts in the World Health Organization World Mental Health Surveys. J Clin Psychiatry. 2010;71(12):1617–1628.

50. Borges G, Angst J, Nock MK, Ruscio AM, Walters EE, Kessler RC. A risk index for 12- month suicide attempts in the National Comorbidity Survey Replication (NCS-R). Psychol Med. 2006;36(12):1747–1757.

51. Mann JJ, John Mann J, Ellis SP, et al. Classification Trees Distinguish Suicide Attempters in Major Psychiatric Disorders. The Journal of Clinical Psychiatry. 2008;69(1):23–31. doi:10.4088/jcp.v69n0104

52. Glenn CR, Nock MK. Improving the short-term prediction of suicidal behavior. Am J Prev Med. 2014;47(3 Suppl 2):S176–S180.

## Supplementary References

1. “A global reference for human genetic variation” Nature 526 68–74 2015

